# Cytomegalovirus infection protects against metastatic melanoma and modulates oncological outcome and toxicity to checkpoint immunotherapy

**DOI:** 10.1101/2024.10.09.24315144

**Authors:** Gusztav Milotay, Martin Little, Robert Watson, Dylan Muldoon, Orion Tong, Chelsea Taylor, Sophie MacKay, Isar Nassiri, Bo Sun, Louisa M Webb, Julia Bremke, Oluwafemi Akin-Adigun, Piyush Kumar Sharma, Weiyu Ye, Ros Cooper, Sara Danielli, Flavia Matos Santo, Alba Verge de Los Aires, James Gilchrist, Esther Ng, Amanda Y Chong, Alex Mentzer, Michael Youdell, Victoria Woodcock, Nicholas Coupe, Miranda J Payne, Paul Klenerman, Mark M Middleton, Benjamin P Fairfax

**Affiliations:** MRC Weatherall Institute of Molecular Medicine, University of Oxford, Oxford, UK; Department of Oncology, University of Oxford, Oxford, UK; Nuffield Department of Clinical Neuroscience, University of Oxford, Oxford, UK; Oxford Cancer — CRUK Oxford Centre,University of Oxford,Churchill Hospital,Oxford.; Department of Paediatrics, University of Oxford, Oxford, UK; Kennedy Institute for Rheumatology, University of Oxford, Oxford, UK; The Centre for Human Genetics, University of Oxford, Oxford, UK; Big Data Institute, Li Ka Shing Centre for Health Information and Discovery, University of Oxford, Oxford, UK; Cancer and Haematology Centre, Oxford University Hospitals NHS Foundation Trust, Oxford, UK; Translational Gastroenterology Unit, John Radcliffe Hospital, Oxford, UK; NIHR Oxford Biomedical Research Centre, Oxford University Hospitals NHS Foundation Trust, Oxford, UK

**Author notes:** Equal contributions. Contributing authors. These authors contributed equally to this work (Gusztav Milotay, Martin Little, Robert Watson).

**Keywords:** Cytomegalovirus, melanoma, Immune Checkpoint Blockade, irAEs, *BRAF*

## Abstract

The relationship between chronic viral infection and cancer response to immune checkpoint blockade (ICB) is poorly understood. Cytomegalovirus (CMV) infection is globally endemic and causes severe disease in the immunocompromised. In immunocompetent individuals the clinical effects of CMV infection are an area of active investigation. Here, in analysis of 396 patients receiving ICB for cancer, we investigate the oncological and immunological consequences of CMV seropositivity. We find that infection with CMV leads to profound skewing of CD8^+^ T cell subsets towards an effector phenotype, divergence in gene expression, increased total lymphocyte count and reduced neutrophil:lymphocyte ratio. These differences are associated with immunologically distinct responses to ICB in patients with metastatic melanoma (MM). We identify a gene set highly-associated with CMV infection which is robustly induced by combination ICB (anti-CTLA-4 + anti-PD-1, cICB) but not by single-agent anti-PD-1 (sICB) in CMV seronegative individuals. Consequently, CMV seropositivity is associated with prolonged Overall Survival (OS) in those treated with sICB (HR 0.49, P.01) whereas there is no observed survival association of CMV following cICB treatment (HR 0.95, P=.82). We demonstrate these CMV-determined divergent effects are driven by expression of *TBX21*, encoding the transcription factor T-bet. Expression of *TBX21* predicts OS across all patients (HR 0.67, P=0.013 for above-median expression), with durable response to ICB associated with induction of expanded *TBX21* expressing CD8^+^ clones. Unexpectedly, we find CMV infection is associated with reduced cumulative incidence of Grade 3+ immune related adverse events (irAEs) at 6 months (0.31 vs. 0.53, P=2.1×10^−5^), notably lowering incidence of colitis (P=.00095) and pneumonitis (P=.026), with infected patients requiring fewer steroids or second line immunosuppressants. Finally we link CMV infection to protection against MM, demonstrating lower seropositivity rates in patients with MM, but not early Stage II/III disease, compared to population controls from the UK biobank (OR 0.53, P=.00016). CMV protection is contingent upon melanoma *BRAF* mutation status, with CMV being associated with reduced development of MM in *BRAF* mutated patients and later presentation of *BRAF* wild-type MM.

This work reveals a previously unrecognised interaction between CMV infection, melanoma mutational state, development of metastatic disease and response to ICB, as well as demonstrating CMV infection protects against ICB irAEs, underpinning the importance of prior infection history and chronic immune activation in development of MM and outcomes to immunotherapy. We anticipate other immunosensitive cancers may show similar interactions between chronic viral infection and response to ICB.

## Background

The majority of tumour infiltrating T cells do not appear to recognize cancer related antigens. Instead, viruses form a major source of putative antigens for these cells which are referred to as ‘bystander’ T cells[1–3]. The degree to which bystander T cells have anti-tumourigenic properties and their role in tumour development across cancer subtypes remains unresolved. One key source of bystander T cell viral antigens is Cytomegalovirus [4] (CMV – Human Herpes Virus 5 – HHV5), a betaherpes virus that causes latent chronic viral infection (CVI) with spontaneous reactivation[5]. Global seroprevalence of CMV infection is high, although seropositivity varies between countries and is associated with sociodemographic factors including population density[6, 7]. In the UK Biobank (UKB) 56.1% of white British aged 40-70 are CMV seropositive, with this rising by approximately 0.7% per year from just under 50% aged 40[8]. Infection with CMV is a cause of serious morbidity in neonates and immunocompromised individuals, whilst, in otherwise healthy people, CMV has been associated with a number of health complications including increased rates of cardiovascular disease and incident mortality[9]. These observations are controversial however due to the potential for confounding, and a recent study in the UKB was unable to demonstrate causal association between CMV and cardiovascular disease[10]. What is incontrovertible though is that CMV infection induces profound changes in T cell immunity. Most notably, CMV elicits the induction of memory T cell inflation. This consists of durable clonal hyper-expansion of effector memory cells re-expressing CD45RA (TEMRA cells) that display preserved effector function[11–13], with consequent skewing of the clonal repertoire[14]. In the context of metastatic melanoma (MM) we have shown that increased numbers of large CD8^+^ T cell clones displaying cytotoxic gene expression profiles both pre and post ICB are associated with improved response to ICB treatment and longer overall survival (OS)[15, 16]. The relationship between CMV infection (which we define as IgG reactive seropositivity) and response to ICB in melanoma has not been exhaustively explored however. Therefore, given bystander clones are often CMV reactive, as well as the correspondence between increased T cell clonality – a correlate of good response to ICB – and CMV infection, we have systematically assessed the relationship between CMV seropositivity and outcomes to ICB for melanoma, analysing data from 365 patients for whom CMV serology could be determined in the Oxford Cancer Immunotherapy Toxicity and Efficacy (OxCITE) cohort study.

### CMV seropositivity and pre-treatment haematological and immune profiles

The total study population was 396 patients within the OxCITE study, with CMV serotyping being performed for 365 patients (median age 68 years, range 18-96 years, IQR 55-75 years) who had received ICB as standard of care treatment for cancer within the NHS. The majority (312/365) of samples were from patients receiving ICB for melanoma (273 MM, 39 adjuvant treatment), with the remainder samples from 53 patients receiving ICB alone for unresectable or metastatic non-melanoma cancer (Table 1 & Table 2 for breakdown). Analysis of pre-treatment hospital derived standard of care blood tests across all patients with metastatic disease demonstrated CMV seropositivity was associated with increased baseline lymphocyte count (n=204, 109 CMV seronegative, median=1.43×10^9^/L, 95 CMV seropositive, median=1.83×10^9^/L, difference in interval –0.35×10^9^/L, 95% CI –0.51:-0.20, P= 1.0×10^−5^ Wilcoxon rank sum test; Fig. 1a). We explored the relationship between CMV serostatus and the neutrophil/lymphocyte ratio (NLR), a predictive marker of treatment response to ICB in melanoma[17], as well as a pan-cancer prognostic marker[18]. In keeping with the negative prognostic association of NLR, we found increasing NLR in MM patients was a hazard for death, with the other key significant clinical factors identified being treatment with sICB and prior treatment of MM with *BRAF* inhibitors (Ext. Fig. S1), known to reduce response to ICB[19]. Interestingly, CMV seropositivity was associated with a significantly reduced NLR (109 CMV seronegative, median= 3.27; 95 CMV seropositive, median= 2.49; interval= 0.78, 95% CI 0.29-1.19, P= 0.0017, Wilcoxon rank sum test, Fig. 1b), linking CMV seropositivity to a favourable pre-treatment immune profile. To develop a more granular picture of the CMV effect in patient samples, flow cytometry was performed on pre-treatment PBMCs where available (n=74), assessing CD4^+^ and CD8^+^ T cell subsets. Consistent with observations in healthy individuals[13] we find CMV seropositivity is associated with highly skewed pre-treatment T cell subsets, with depletion of naive and central memory subsets and expansion of effector memory and TEMRA subsets (Fig. 1c, Fig. 1d). Given their key role in the response to ICB we focused on CD8^+^ T cells, exploring the pre-treatment effect of CMV serostatus across patients with MM for whom expression and CMV serology data were available. Analysis of the effect of CMV serostatus on CD8^+^ T cell gene expression in baseline samples from 186 patients (82 CMV seropositive, 104 CMV seronegative) demonstrated CMV to be associated with profound divergence in CD8^+^ pre-treatment expression, with differential expression of 7,175/17,440 transcripts detected (P_adj._*<*0.05, 19% of all transcripts upregulated, 22% downregulated; Fig. 1e, Supplementary Table 1). Gene ontology analysis indicated specific regulation of 146 pathways (109 induced, 37 downregulated, Fig. 1f, Supplementary Table 2) involving key CD8^+^ T cell functions – notably CMV seropositivity was associated with upregulation of T Cell Receptor signalling (P_adj._= 3.7×10^−5^), Interferon *γ* mediated signalling (P_adj._=3.7×10^−5^) and Antigen processing and presentation via Class I MHC (P_adj._= 1.0×10^−4^), all of high importance to anti-cancer immunity. Many of the CMV upregulated genes encoded cell surface proteins implicated in T-cell signalling including *CD2*, previously noted to be associated with with enhanced CAR-T tumour killing [20], *LAG3*, the target of the immunotherapy relatlimab[21], and *CD8A*, a T cell co-receptor. We additionally observed upregulation of the transcription factor *ZNF683*, implicated in T cell response to recurrent antigen exposure and identified to show specific expression in HCMV-reactive T cells[22, 23], and latterly implicated in resident memory populations involved in CD8^+^ T cell anti-PD1 immunotherapy induced anti-tumour responses[24].

**Fig. 1.**
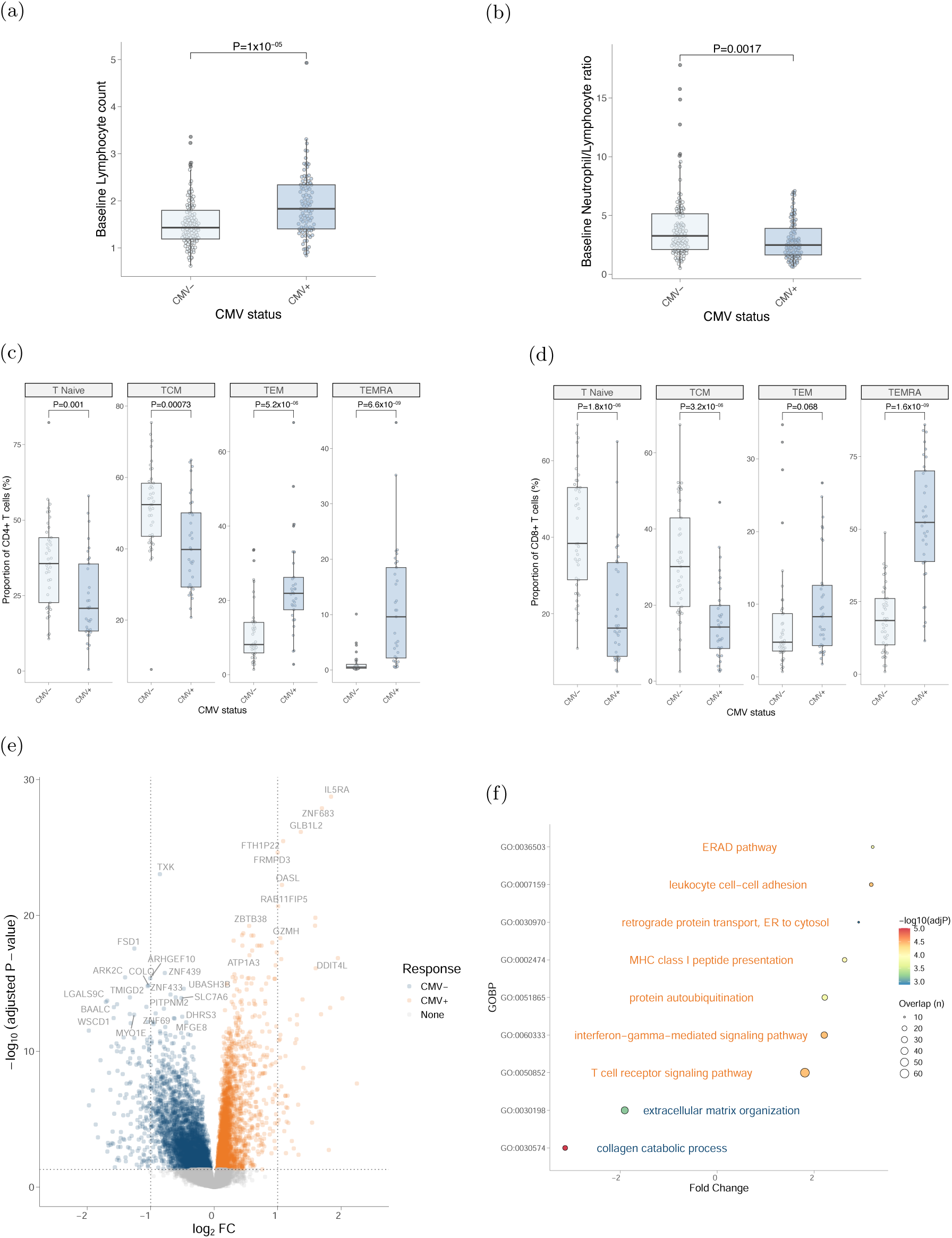
(a) Pre-treatment lymphocyte count is elevated in CMV seropositive metastatic melanoma patients (b) CMV pre-treatment neutrophil:lymphocyte ratio is reduced in CMV seropositive patients (c) Flow cytometry derived pre-treatment CD4^+^ T cell subset proportion according to CMV serostatus indicating expansion of TEMRA / effector memory subsets (d) As per (c) but for CD8^+^ subsets (e) Differentially expressed genes (DEG) according to CMV serostatus from pre-treatment CD8^+^ T cells, y-axis shows –log10(P_adj._) derived from Wald test using CMV seronegative samples as a reference (f) Gene Ontology Biological Process (GOBP) analysis of DEG from (e) highlighting induction/suppression (orange and blue respectively) of pathways involved in T cell activation in CMV seropositivity. All tests Wilcoxon rank sum test unless otherwise stated.

**(a) Table 1.**
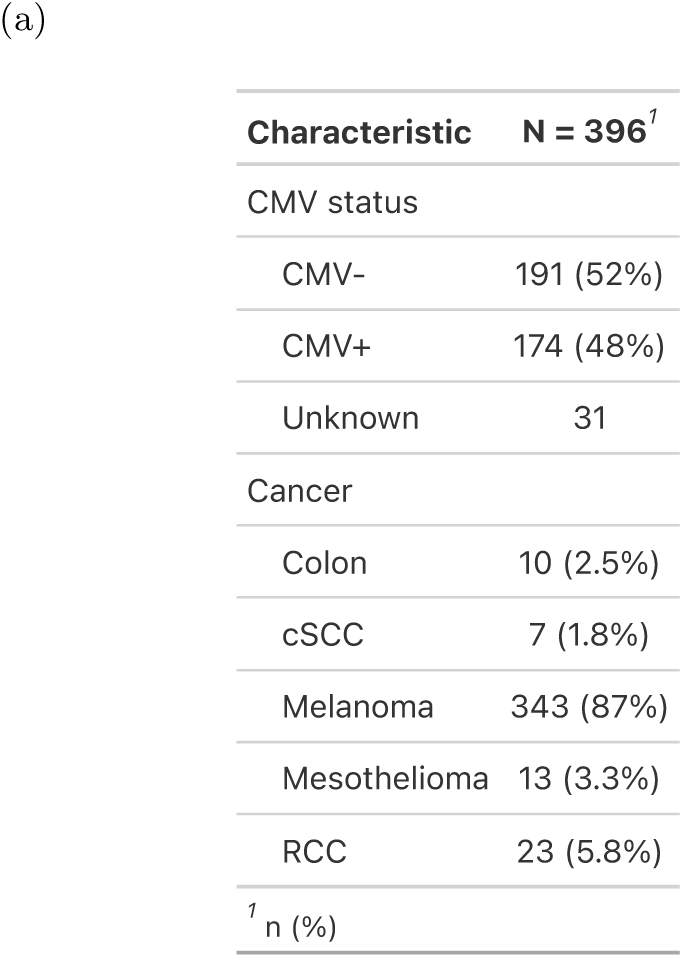
– Breakdown of all patients by cancer type within the OxCITE cohort.

| Characteristic | N = 396<sup>†</sup> |
| --- | --- |
| CMV status |  |
| CMV- | 191 (52%) |
| CMV+ | 174 (48%) |
| Unknown | 31 |
| Cancer |  |
| Colon | 10 (2.5%) |
| cSCC | 7 (1.8%) |
| Melanoma | 343 (87%) |
| Mesothelioma | 13 (3.3%) |
| RCC | 23 (5.8%) |
| <sup>†</sup> n (%) |  |

**(b) Table 2.** – Breakdown of patients within the OxCITE cohort with melanoma.

| Characteristic | N = 343<sup>†</sup> |
| --- | --- |
| BRAF status |  |
| Mutant | 116 (34%) |
| Wild-type | 221 (66%) |
| Unknown | 6 |
| CMV status |  |
| CMV- | 162 (52%) |
| CMV+ | 150 (48%) |
| Unknown | 31 |
| Age | 67 (55, 75) |
| Sex |  |
| Female | 154 (45%) |
| Male | 189 (55%) |
| Treatment |  |
| AdjNivo | 9 (2.6%) |
| AdjPembro | 30 (8.7%) |
| Ipilimumab | 2 (0.6%) |
| IpiNivo | 195 (57%) |
| Nivo | 16 (4.7%) |
| Pembro | 88 (26%) |
| RelatNivo | 3 (0.9%) |
| <sup>†</sup> n (%); Median (IQR) |  |

### CMV serostatus is associated with transcriptomic and clinical response to ICB

Given the favourable association of CMV seropositivity with pre-treatment haematological indices and the divergence in baseline CD8^+^ T cell gene expression according to CMV status, we explored the effect of ICB treatment on CMV-associated gene expression. To capture CMV-associated genes in one value we developed a CMV transcriptomic score based on the first principal component of genes associated with CMV serostatus, incorporating the number of CMV-associated genes that provided a maximal AUC for CMV serostatus prediction from CD8^+^ T cell transcriptomics alone (103 genes providing an AUC of 0.9, Ext. Fig. S2a). Correspondingly, pre-treatment samples showed a highly significant difference in CMV score according to serostatus (Fig. 2a). The CMV score was associated with the proportional composition of TCR within the transcriptome – a surrogate for clonal size – with a significant positive correlation between CMV score and the number of TCR comprising *>*0.5% the clonal repertoire, which we have previously shown to be prognostic both pre and post ICB [15, 16] (Ext. Fig. S2b, Fig. 2b). Given the CD8^+^ transcriptional response significantly differs between sICB and cICB treatments [15] we assessed each treatment individually. In CMV seropositive patients we observed no additional effect of either treatment on the CMV score. Conversely, in CMV seronegative patients the response varied according to ICB given, with patients receiving cICB demonstrating an increase in CMV score (P=6.4×10^−9^, Wilcoxon signed rank test). This was not seen in sICB treatment however with no significant change in CMV score in either patient group (Fig. 2c). A similar effect was noted when the expression of cytotoxic genes was assessed – corresponding to a previously described prognostic cytotoxicity score [16] (Fig. 2d, Supplementary Table 3). This analysis demonstrates cICB treatment leads to partial transcriptomic convergence of CD8^+^ T cell gene expression between CMV seropositive and seronegative patients, whereas no effect is noted with anti-PD1 alone, indicating the immunological response to ICB in MM depends both upon the CMV status of the patient and the addition of anti-CTLA-4 treatment. To explore clinical implications of these observations, Kaplan-Meier (KM) analysis of OS in patients with a minimum of 6 months follow-up post commencement of ICB was performed. Within the cICB treated patients we found no significant difference in median OS according to CMV serostatus (n=181, median survival 107 CMV seronegative:78.2 months, 74 CMV seropositive:60.5 months, HR=0.95, P=0.82, Log-rank test, Fig. 2e). Similar results were observed when other variables were taken into account in a multivariable Cox proportional hazards model (Fig. 2f). Analysis of sICB treated patients however demonstrated that CMV seropositive patients had significantly prolonged OS compared to CMV seronegative patients (n=72, 28 CMV seronegative, median OS 34.4 months; 44 CMV seropositive, median OS not reached, HR=0.49, P=0.037, Log-rank test, Fig. 2g) which was robust to multivariable analysis (P= 0.0085, Fig. 2h). Notably multivariable analysis also highlighted that upon controlling for CMV, age becomes a significant risk factor for death (P=0.0079), in keeping with evidence that the effects of CMV are distinct to those found in age-associated immunosenescence[25]. In sum, these results support CMV seropositivity conferring induction of a prognostically favourable CD8^+^ T cell gene set that is only invoked by combination CTLA-4/PD-1 blockade, resulting in divergent outcomes according to CMV serostatus in recipients of single agent PD-1 blockade.

**Fig. 2.**
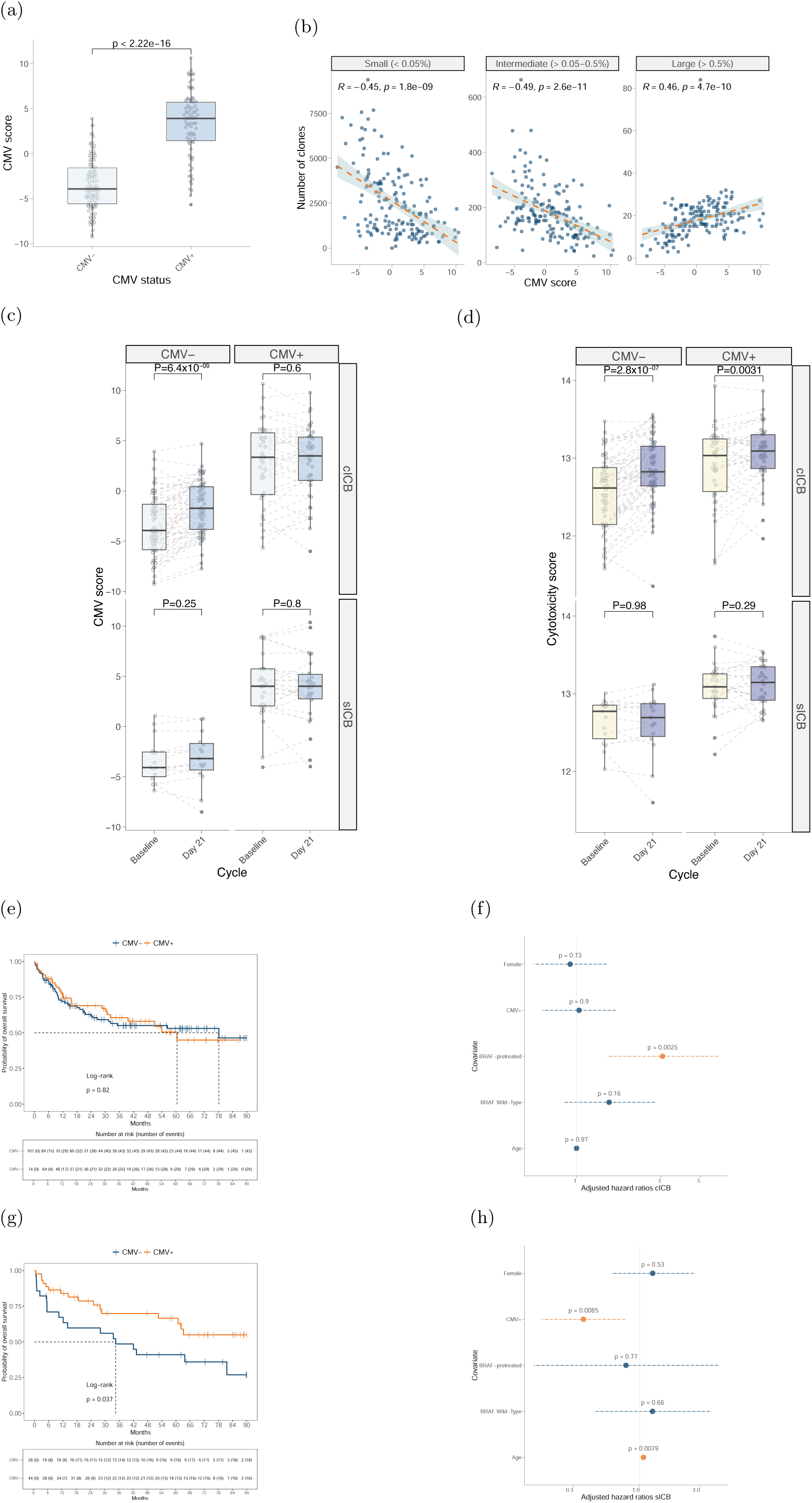
(a) CMV score of pre-treatment patient CD8^+^ T cell samples according to CMV seropositivity (Wilcoxon rank sum test) (b) CD8^+^ T cell clones, stratified according to size (facets) and number (y axis), versus CMV score post one cycle of immunotherapy demonstrating a skewing of the repertoire towards expanded clones in patients with a increasing CMV score (Spearman’s Rank Correlation test) (c) cICB treatment of CMV seronegative patients leads to a highly significant induction of CMV score (y axis) (top left panel), not observed in CMV seronegative patients receiving sICB (bottom left panel). No significant induction of CMV score is noted in CMV seropositive in either cICB (top right panel) or sICB recipients (bottom right panel). P values derived from Wilcoxon signed rank test (d) cICB treatment leads to a significant induction of cytotoxicity score irrespective of CMV status (y axis) (top panels), not observed in patients receiving sICB (bottom panels). P values derived from paired Wilcoxon test (e) Kaplan Meier analysis shows no significant difference in OS of cICB patients between CMV seropositive and CMV seronegative patients (P=0.82, log-rank test) (f) Cox proportional hazards model controlling for age, sex, *BRAF* mutation status and prior treatment with *BRAF* inhibitors shows no significant hazard ratio for death with CMV serostatus in cICB recipients (g) Kaplan Meier analysis demonstrates increased OS of CMV seropositive versus seronegative sICB recipients (P=0.037, log-rank test) (h) Cox proportional hazards model as per (f) shows significantly reduced adjusted hazard ratio for death in CMV seropositive recipients of sICB. Increased age is significantly associated with increased risk of death when controlling for CMV.

### CMV seropositive patients have fewer severe immune related adverse events

Immune related adverse events (irAEs) are a frequent complication of ICB treatment, with Grade 3 or above irAEs, which represent the most clinically significant toxicities, observed in real-world clinical data in 10-15% of sICB recipients and 60~% of cICB recipients[26–28]. Whilst irAEs are a cause of morbidity and increased demand on healthcare resources, multiple studies have reported associations between the development of irAEs and improved survival in patients receiving ICB for both MM and other cancers[26, 29, 30]. We assessed the relationship between the development of irAEs and clinical outcome in ICB treated MM using a time-dependent Cox-regression model to account for guarantee-time bias susceptibility[31]. Analysis at the level of organ-specific irAEs demonstrated that, in keeping with other studies, both arthritis and dermatitis were associated with improved OS[32]. Summarising across irAEs showed that mild Grade 1-2 irAEs were significantly associated with improved survival, whereas Grade 3+ irAEs were not (Ext. Fig. S3a). This is in keeping with recent meta-analyses and may reflect that higher grade irAEs typically require treatment with immunosuppressants including steroids, potentially antagonising the anti-cancer effect of ICB[29, 33, 34]. Given our findings, we were interested to see whether CMV influenced irAE development. We estimated cumulative incidence and time to irAEs using a univariate semi-competing risks model (death being the competing risk) across all melanoma patients treated with anti-PD-1 or anti-PD-1/anti-CTLA-4 (n=307). Whilst no effect of CMV seropositivity was seen with Grade 1-2 irAEs (Ext. Fig. S3b), CMV was associated with delayed time to onset of severe Grade 3+ irAEs and a significantly reduced cumulative incidence of for Grade 3+ irAEs (0.53 vs 0.31 at 6 months, P= 2.1× 10^−5^, Fig. 3a). We further explored this using multivariable analysis controlling for age, sex, ICB type, *BRAF* status and prior *BRAF* inhibitor treatment. This demonstrated that, when taking account of these variables, both the known comparative reduced toxicity of sICB and CMV remained independently associated with significantly reduced incidence Grade 3+ irAEs (P= 0.0056, Fig. 3b). We then analysed this CMV association across the entire time-course of patient ICB treatment using binomial logistic regression, similarly noting no association with Grade 1-2 irAEs, but a significantly reduced incidence of Grade 3+ irAEs (OR=0.49, P=7.8×10^−5^, Fig. 3c). In keeping with this, we found patients with CMV had strikingly reduced requirements for immunosuppression both in the form of steroids (OR=0.67, P=0.030) and second-line agents given upon steroid failure to control toxicity (OR=0.41, P=0.0030). To understand whether CMV leads to reductions across all irAEs, or has organ-specific effects, we explored irAE affected organs versus CMV serostatus across all patients (Fig. 3d). This showed CMV seropositivity is specifically associated with reduced incidence of distinct irAEs, these being colitis (P=0.00095, OR=0.40), pneumonitis (P=0.026, OR=0.23) and myalgia (P=0.012, OR=0.12). We also noted CMV conversely leads to a significantly increased risk of skin irAEs (P=0.049, OR=1.64), but could detect no other discernible effect on any other organ specific irAEs.

**Fig. 3.**
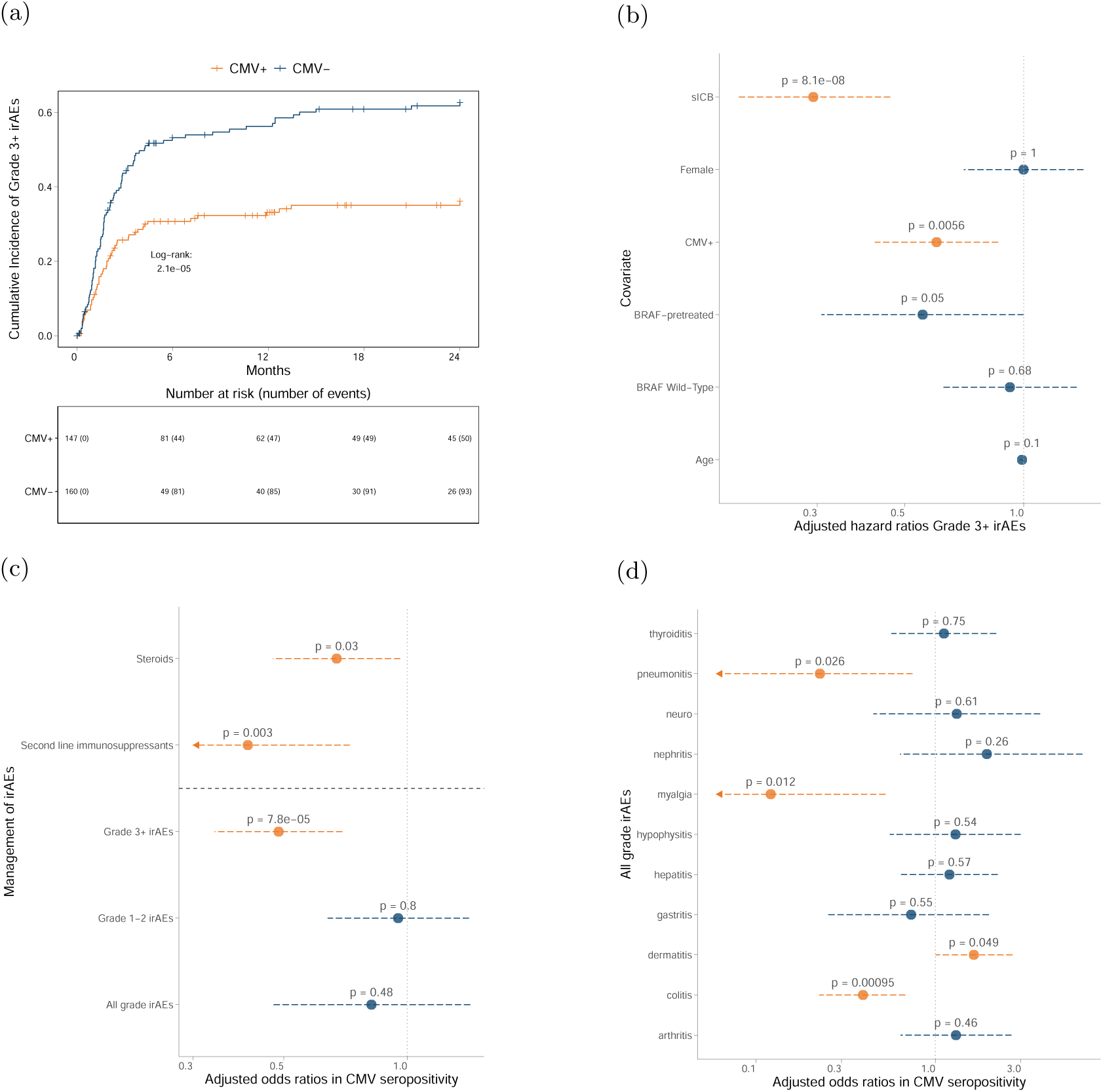
(a) Cumulative incidence of developing Grade 3+ irAEs in all ICB treated melanoma patients is significantly reduced in CMV seropositivity (P=2.1×10^−5^, log-rank test) (b) Multivariate semi-competing risks analysis, where death is a semicompeting risk, indicates that CMV is protective against Grade 3+ irAE development when adjusting for age, sex, treatment type, *BRAF* mutation status and prior *BRAF* inhibition as covariates (c) Time independent adjusted odds ratios for development of all grade, grade 3+, grade 1-2 irAEs and requirement of steroid or second line immunosuppression in CMV seropositive patients, adjusting for covariates as per (b) (Binary logistic regression)(d) Adjusted odds ratios for development of all grade organ specific irAEs in CMV seropositive patients (Binary logistic regression) adjusting for covariates as per (b)

### *TBX21* drives CMV associated CD8^+^ T cell gene expression

The high conservation of the CMV associated transcriptional profile across seropositive patients suggested it could be driven by specific transcription factors (TFs). To identify such TFs we interrogated the output from CMV CD8^+^ T cell differential expression analysis with decoupleR [35], which infers regulatory TFs from specific gene sets. This highlighted 46/792 CD8^+^ T cell expressed TFs demonstrating nominal association with CMV regulated genes (Fig. 4a, Supplementary Table 4), 36 of which had induced activity in CMV seropositivity (P*<*0.05, Univariate Linear Model) and 13 of which were significantly upregulated in CD8^+^ T cells from CMV seropositive patients (Ext. Fig. S4a). We independently correlated the CMV score with expression of each of the 36 induced TFs, finding *TBX21*, encoding T-bet, to be the most strongly associated and thus most plausible causal TF (rho=0.76, P*<*2.2×10^−16^, Fig. 4b, Ext. Fig. S4b). This observation fits with the known immunological function of T-bet, a key determinant of CD8^+^ T cell effector activity [36] that is associated with the development of terminally differentiated anti-viral CD8^+^ T cells with comparatively low expression of PD-1[37]. We subsequently explored the expression of *TBX21* across MM patient CD8^+^ T cell bulk RNA-seq data, both prior to and post ICB. Flow cytometry based analysis of pre-treatment CD8^+^ cell subsets showed a strong relationship between total *TBX21* expression and the subset composition per patient, with *TBX21* correlating with proportion of CD8^+^ TEMRA cells, and highly anti-correlated with CD8^+^ Naive cells (Fig. 4c). *TBX21* expression displayed a similar relationship with clone size to that of the CMV score and, consistent with the crucial role of *TBX21* in memory inflation[38], was correlated with increasing number of post-ICB large clones (rho= 0.52, P= 8.5×10^−13^, Ext. Fig. S4c). In keeping with *TBX21* driving much of the CMV associated signature in these cells, *TBX21* expression in response to ICB was similar to the CMV score – with increased baseline expression of *TBX21* in CMV seropositive individuals, whilst *TBX21* induction was only observed in CMV seronegative patients receiving cICB (P= 0.0026, Wilcoxon signed rank test, Fig. 4d). Interestingly, sICB treatment was associated with a small but significant reduction in *TBX21* expression in CMV seronegative individuals (P= 0.0026). Similarly, although subset composition remained stable according to CMV status across treatment, CD8^+^ GZMB^+^ T cells were observed to significantly increase in cICB treated CMV seronegative patients, whereas there was a fall in this population in CMV seronegative patients receiving sICB (Fig. 4e). Granzyme B is under the control of *TBX21* [39], and these independent flow cyometry observations corroborate the transcriptomic analysis. To further refine observations made from bulk CD8^+^ T cell RNA-seq we performed analysis at the single cell (sc) level, exploring the relationship between *TBX21* expression, CD8^+^ T cell subsets and individual T cell clones. We generated sc-RNAseq data for 18 patients with MM pre and post sICB treatment (median 1919 cells per sample baseline/ 1941 cells day 21). *TBX21* was robustly expressed in CD8^+^ T effector (T_Eff_) cells – corresponding to the flow cytometry TEMRA subset, consistent with prior descriptions[36], as well as *γδ* T cells (T*_γδ_*), Effector Memory (T_EM_) populations, and to a lesser extent, MAIT cells (Fig. 4f). We therefore focused upon the T_Eff_ subset which had the highest median *TBX21* expression. Given pathway analysis of CMV associated expression profiles in bulk RNAseq demonstrated upregulation of TCR signaling (Fig. 1f), we assessed the number of TCR *β* chains sequenced per cell, as defined by the unique molecular identifiers (UMI), comparing those from cells with and without detected expression of *TBX21*. This showed that cells with detectable *TBX21* expression have increased TCR *β* chain numbers both pre and post sICB treatment (Fig. 4g), indicating a direct relationship at the individual cell level between *TBX21* expression and enhanced TCR signaling. Conserved clonal patterns of gene expression are indicative of shared antigen target and functionality, with clonal expansion reflecting antigen recognition. Further supporting a key role for *TBX21* expression we found large clones (those *>*0.5% total repertoire) had significantly increased *TBX21* expression post-treatment (Fig. 4h), reinforcing the relationship between CMV, *TBX21* and clonal expansion.

**Fig. 4.**
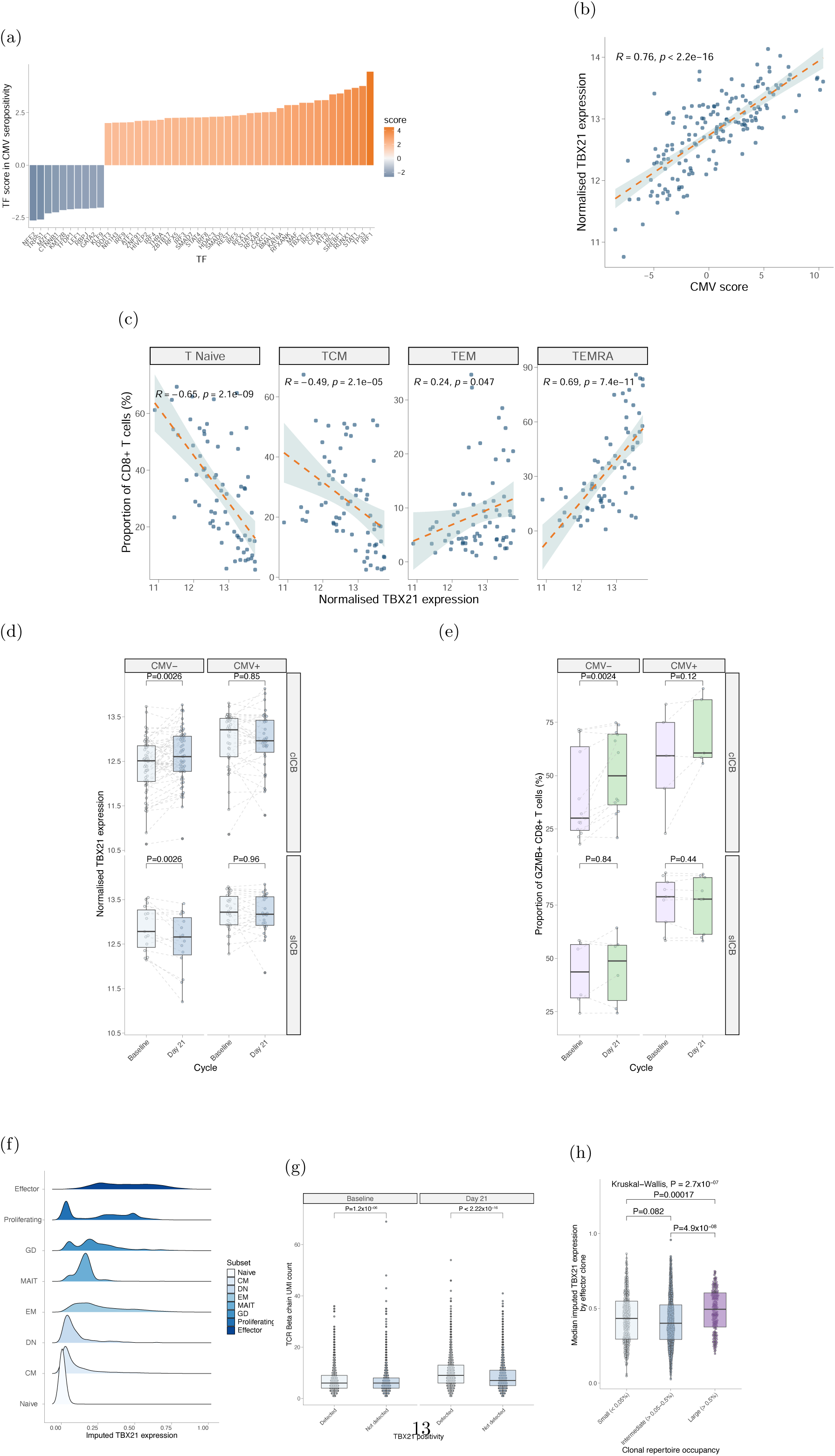
(a) Transcription factor activity inference in CMV seropositive baseline CD8^+^ samples (b) Normalised CD8^+^ *TBX21* expression correlated with CMV score post one cycle of immunotherapy (Spearman’s Rank Correlation test) (c) Baseline CD8^+^ subset correlations with *TBX21* highlights that increased *TBX21* expression is correlated with increased circulating TEMRA and decreased Naive T cell proportions (Spearman Rank test) (d) In CMV seronegative patients, treatment with cICB (top left panel) is associated with a significant induction of *TBX21*. This is not observed in CMV seronegative patients receiving sICB (bottom left panel) where *TBX21* is significantly downregulated. In CMV seropositive patients no significant change in *TBX21* expression is noted in either cICB recipients (top right panel) or sICB recipients (bottom right panel). All P values result from application of a Wilcoxon signed rank test (e) Flow cytometry blinded to CMV and ICB type measuring the proportion of GZMB^+^ CD8^+^ T cells pre and post ICB, dichotomised by CMV status and ICB type. P values relate to Wilcoxon signed rank test. (f) Imputed *TBX21* expression by CD8^+^ subset in sICB-treated patients Relative expression of *TBX21* by CD8^+^ subset as assessed using scRNA-seq (g) TCR *TRB* UMI count per effector cell stratified by *TBX21* detection pre and post-immunotherapy (Wilcoxon rank sum test) (h) Median imputed *TBX21* expression by effector clone stratified by peripheral repertoire occupancy in post-treatment samples, demonstrating that *TBX21* is a marker of clonal expansion (Kruskal-Wallis test)

### CD8^+^ T cell *TBX21* expression is independently associated with durable response to ICB

We reasoned that if the relationship between CMV infection and outcome from ICB is mediated by expression of *TBX21*, then CD8^+^ T cell *TBX21* expression may predict response to ICB. Analysis of all patients within the scRNA-seq dataset demonstrated that clones from patients with a durable response to ICB (defined as absence of progression within 3 years of treatment initiation) displayed a pronounced induction of *TBX21* expression upon initiation of ICB treatment, whereas clones from patients with progressive disease did not show this response, instead showing relatively high pre-treatment *TBX21* expression with significant reductions in overall *TBX21* clonal expression upon ICB treatment (Fig. 5a). We further assessed the relationship between *TBX21* expression and outcome from CD8^+^ T cell bulk RNA-seq dataset over three year patient outcomes, finding baseline differences in median *TBX21* expression according to outcome that became more apparent after one cycle of ICB (Fig. 5b). We proceeded to formally assess the prognostic relationship between posttreatment CD8^+^ T cell *TBX21* expression and clinical outcome using univariate KM analysis across all ICB treated MM patients for whom we had a post-treatment day 21 sample (n=164). This showed a highly significant difference in progression free survival (PFS) based on *TBX21* expression alone, with patients with below median CD8^+^ T cell *TBX21* expression having a median PFS of 7.7 months versus 36.5 months for those with above median *TBX21* expression (HR=0.67, P= 0.0020, Log-rank test, Fig. 5c), with this difference extending to OS (HR=0.67, P= 0.013, Log-rank test, Ext. Fig. S5a). Finally we assessed the relationship between CD8^+^ T cell *TBX21* expression using a multivariable analysis, controlling for age, sex, ICB type, *BRAF* status and pre-treatment with *BRAF* inhibitors. This demonstrated that CD8^+^ T cell *TBX21* expression was the crucial predictor of both PFS (Ext. Fig. S5b) and OS (Fig. 5d), with no other covariates showing association with outcome except increasing age which, once *TBX21* expression had been controlled for, showed a negative association with OS (P= 0.030).

**Fig. 5.**
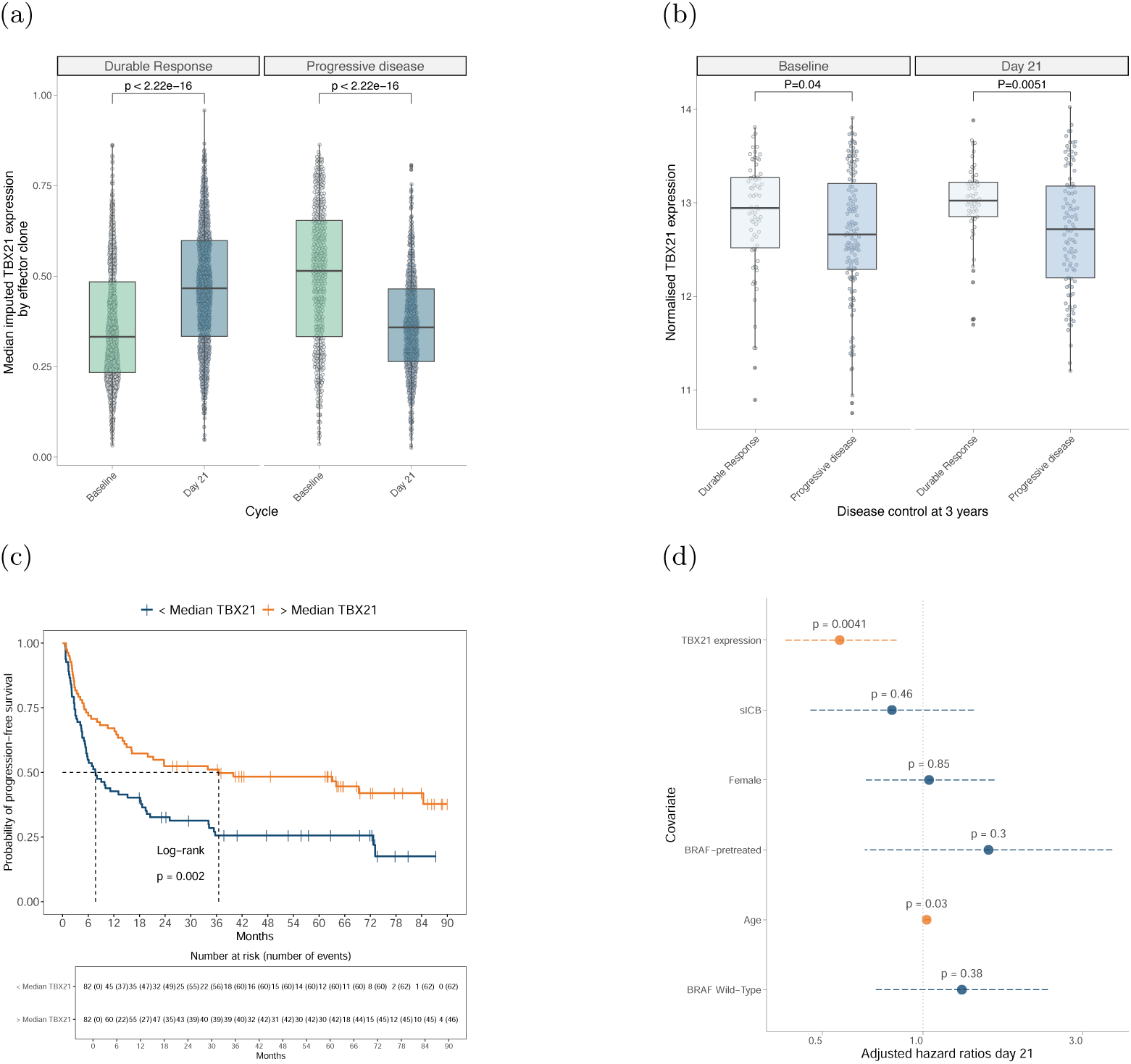
(a) Median imputed *TBX21* expression in effector clones, stratified by disease control 3 years post-initiation of systemic treatment for metastatic disease, highlighting highly significant induction of *TBX21* in patients who respond to treatment while a significant downregulation is noted in patients who progress (b) Bulk CD8^+^ *TBX21* according to disease control at 3 years post-initiation of systemic treatment (c) Kaplan Meier analysis demonstrates significantly improved in PFS in patients with above median *TBX21* post one cycle of immunotherapy (P=0.0020, log-rank test) (d) Cox proportional hazards model indicates that increased post-treatment *TBX21* is associated with improved OS when adjusting for ICB type, age, sex, *BRAF* mutation status and prior *BRAF* inhibition. Increased age is associated with increased risk of death when controlling for *TBX21*

### CMV seropositivity influences metastatic melanoma epidemiology

In view of the interaction between CMV and ICB response we questioned whether CMV might also act earlier to influence the development of MM. Comparative analysis of CMV seropositivity within the UKB (n=7885, white British, age 40-70) to OxCITE cohort patients in the same age group identified a highly significantly reduced rate of CMV seropositivity in samples from MM patients (rate MM 60/149 – 40%, rate UKB 4425/7885 – 56%, OR=0.53, P= 0.00016). Conversely, the CMV seropositive rate in OxCITE cohort patients receiving adjuvant ICB for Stage II/III melanoma was not significantly different from the UKB. The reduced CMV seroprevalence in the MM group was observed across both patients with *BRAF* mutated and wild-type (WT) disease, although the effect was larger in *BRAF* mutated (*BRAF* mutated: OR=0.44, P=0.0018, *BRAF* WT: OR=0.58, P=0.012, Fig. 6a). A restricted permutation analysis to all MM with matched age and sex pairing with UKB samples produced a similar observation (median P= 0.0076, range: 0.0051-0.011, Fisher-Pitman test), whilst subgroup analysis against data from individual geographically disparate UKB collection centres demonstrated a consistent direction of effect across all centres with the complete UKB cohort, ruling out a sociodemographic effect (Ext. Fig. S6a). In support of an increased sensitivity of *BRAF* mutated melanoma to CMV, analysis across all OxCITE patients with MM demonstrated that there were fewer CMV seropositive *BRAF* mutated patients than expected (33/95 vs. 83/156, P= 0.0060, Fisher’s exact test vs *BRAF* WT, Fig. 6b). Finally, we observed that CMV seropositive patients patients initiating ICB for MM were on average older (65 yr CMV seronegative vs. 70.5 yrs seropositive – median 4 yr difference, 95% CI 2.0-9.0, P= 0.0020, Wilcoxon rank sum test), an effect not observed in other cancers receiving ICB (Fig. 6c). Notably however, when we explored whether this was *BRAF* mutation specific there was no difference in the age of ICB initiation for *BRAF* mutated patients between seropositive and seronegative patients, irrespective of pre-treatment with *BRAF* inhibition (P=0.56). Conversely, *BRAF* WT patients CMV seropositive patients presenting with metastatic disease were a mean 8.0 years older than CMV seronegative *BRAF* WT patients (95% CI 3.0-12.0 years, P= 0.00043, Wilcoxon rank sum test, Fig. 6d). Formal testing supported an interaction with age of presentation according to CMV seropositivity and *BRAF* mutation status (P= 0.014, ANOVA), suggesting that the described later presentation of *BRAF* WT metastatic disease[40] may in part be due to an unrecognised interaction with CMV status. When CMV seropositive patients are excluded we observe no significant difference in age of presentation with metastatic disease across all patients between *BRAF* mutated and WT patients (P= 0.21, Wilcoxon rank sum test). To further understand where in the development of MM CMV was acting in *BRAF* WT melanoma we identified the age of first presentation of primary melanoma for 159 patients. This demonstrated that CMV did not appear to influence the time from primary diagnosis to metastatic disease, but was associated with a significant difference in the age of diagnosis of primary *BRAF* WT melanoma (Ext. Fig. S6b). Given a proportion of the patients would have seroconverted post diagnosis of primary melanoma, this difference is likely an underestimate.

**Fig. 6.**
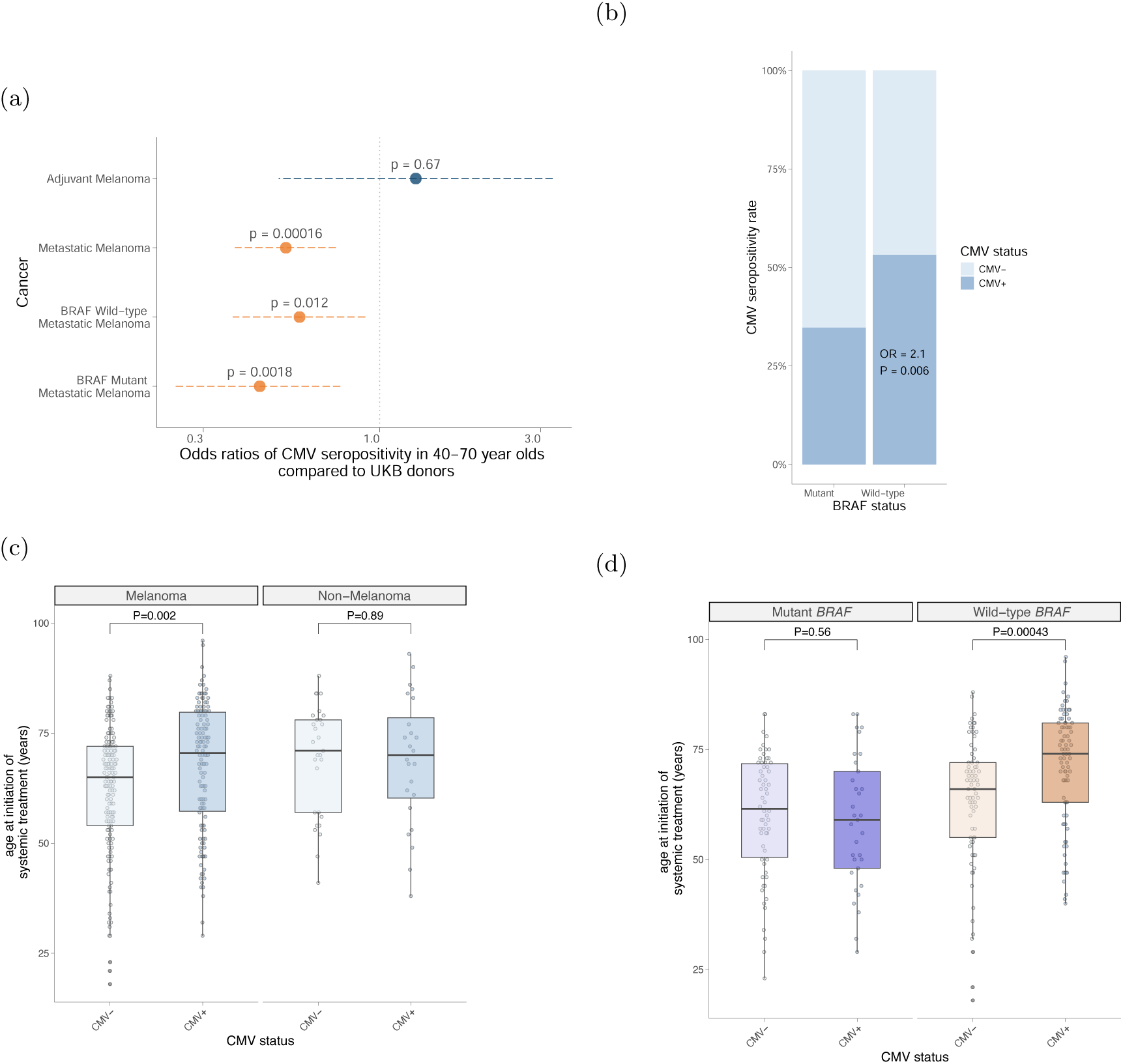
(a) CMV seropositivity of patients with MM and Stage II/III (age 40-70) versus those rates within UKB in white British aged 40-70 (b) Breakdown of *BRAF* mutation status versus CMV serostatus across all patients with MM in the cohort (Fisher’s exact test) (c) Presentation with MM occurs significantly later in patients seropositive for CMV (left panel, 65 yr CMV seronegative vs. 70.5 yrs CMV seropositive, Wilcoxon rank sum test), whilst no significant difference is noted in other ICB-treated metastatic cancer (right panel)(d) Relationship between age of presentation with MM and *BRAF* V600 mutation status according to CMV serostatus (left panel: *BRAF* V600 mutated, right panel *BRAF WT*). There is no significant difference in age of presentation of *BRAF* mutated patients according to CMV serostatus, whereas a highly significant effect is noted in patients with *BRAF* WT MM (right panel, 66 yrs CMV seronegative vs. 74 yrs CMV seropositive, Wilcoxon rank sum test)

## Discussion

In an analysis of immunophenotyping, CD8^+^ T cell transcriptomics and clinical outcome data from 396 patients in the OxCITE cohort, we have performed the first comprehensive study of the effects of CMV on immunological and clinical response to ICB in melanoma. We find in metastatic melanoma that CMV is associated with favourable pre-treatment haematological markers of response to ICB, namely reduced NLR, increased T cell clonality, and higher cytotoxicity, and that CMV causes a distinct pan CD8^+^ T cell transcriptomic profile, with effector skewing of CD8^+^ subsets. We find that combination but not single-agent ICB leads to induction of CMV-associated gene expression in CMV seronegative individuals, resulting in indistinguishable outcomes between CMV seropositive and negative cICB recipients only. We also find that CMV seropositivity is protective against the development of high-grade irAEs, with CMV positive patients requiring less corticosteroids and second-line immunosuppression. We determine that the effects of CMV seropositivity can at least in part be explained by increased expression of *TBX21*, encoding the transcription factor T-bet, which we show independently associates with improved OS following ICB treatment for MM. Finally, we demonstrate CMV seropositivity is protective against the development of MM, with a significantly lower proportion of MM patients seropositive for CMV compared to earlier stage patients and the background population. We further describe a novel and entirely unanticipated interaction between CMV and melanoma *BRAF* mutation status, with CMV being associated with reduced development of MM in *BRAF* mutated patients, whilst primarily leading to delayed onset of *BRAF* WT melanoma. Given the key role of CD8^+^ T cells in the response to ICB in MM, it is perhaps not surprising that the striking divergence in gene expression among CD8^+^ T cells on the basis of CMV serostatus is associated with variation in response and outcome to ICB. CMV seropositive patients have marked upregulation of a number of T cell activatory pathways including T Cell Receptor signalling, IFN*γ* mediated signalling and antigen processing and presentation via Class I MHC. Their T cell subsets are skewed with greater proportions of T_Eff_ and T_EM_ cells. *TBX21* is central to these changes with cells expressing *TBX21* demonstrating upregulated TCR *β* chains in single-cell analysis. Indeed, *TBX21* expression alone is a significant predictor of survival following ICB. Importantly, CMV seronegative individuals can gain a CMV seropositive-like CD8^+^ gene expression profile through treatment with cICB, but not sICB. This has cardinal implications for clinical practice. Treatment with cICB has a small but significant improved OS benefit to sICB at the cost of a marked increased risk of high-grade irAEs[41]. Further, a substantial proportion of patients gain long-term disease control with sICB alone and the identification of these patients is of high clinical importance. Thus CMV serostatus may allow personalisation and stratification of therapeutic approaches, with CMV seronegative patients gaining a clear immunotranscriptomic benefit from cICB, but not sICB.

Perhaps counter-intuitively we find CMV infection is highly protective against Grade 3+ irAEs, particularly colitis and pneumonitis. Conversely, CMV does not protect against lower grade irAEs which tend not to require immunosuppression, but are most strongly linked to improved clinical outcomes. Similarly, CMV is associated with increased rates of cutaneous irAEs, which tend not to require systemic immunosuppression and are prognostically favourable in MM[29, 33]. Both the lungs and the large intestine are frequent sites of CMV reactivation in immunocompromised individuals[42–44] and our observations may reflect that sites of high CMV infection are particularly sensitive to the immuno-modulatory effects of CMV, including the development of tissue resident memory cells [38, 45]. Notably, whilst EBV has recently been directly linked to development of multiple sclerosis (MS), the same study demonstrated CMV infection was protective against MS development[46], replicating an earlier observation[47]. Thus, it is clear that different CVIs can have pleiotropic organ specific effects on inflammatory and autoimmune conditions. Similarly, irAEs have been associated with increased TCR diversity [48] which corresponds to T cell naivety – the observed CMV associated reduction naive subsets could therefore be a further mechanism of protection. Dissection of this effect will require in-depth immunophenotyping of the relevant organs, taking into account CMV serostatus, and, whilst outside the scope of this original report, is of high importance to better understanding ICB toxicity.

Finally, the transition from localised to metastatic disease involves a myriad of tumour:host interactions that influence the probability of tumour dissemination. Crucially, metastasising cells must evade T cell immunity to develop distal disease. By testing against the UKB – the largest demographically relevant population control cohort – we detected a highly significantly reduced rate of seropositivity in MM but not Stage II/III melanoma. Together these observations would fit with a model whereby infection with CMV invokes a CD8^+^ T cell profile that has anti-melanoma properties – manifesting as protection against onset of metastatic disease. Perhaps most striking however is that this CMV effect diverges according to melanoma mutation status, with significantly fewer *BRAF* mutated CMV positive MM patients than expected. Moreover, CMV delays but does not prevent development of *BRAF* WT MM. Given ICB instigates a prominent anti-viral response[15], it is not surprising that the antiviral response towards CMV confers anti-tumour activity. *BRAF* mutated melanomas generate an immune response distinct to that of *BRAF* WT tumours, with reduced CD8^+^ T cell infiltration[49] and improved outcomes to cICB[41]. Therefore, *BRAF* mutated tumours appear to show sensitivity to a preventative effect of CMV, whereas we determine the development of *BRAF* WT MM to be delayed by years, albeit not prevented.

We note the limitations of the current study. The data is observational and as such, caution must be applied when trying to establish causation. Our analysis focuses entirely on human data and this limits the ability to generate a causal functional model. Nonetheless, our utilisation of a very large cohort of patients receiving ICB within a standard of care setting, in conjunction with a large population relevant control dataset, means our findings are pertinent to clinical practice. We have a high dimensional dataset for each participant including annotated follow up, transcriptional profiling, CMV serotyping and single cell and flow cytometry analysis on large subsets. Given the almost equal distribution of CMV carriers and non-carriers in the UK, the cohort is powered to detect substantial clinically meaningful differences with a high degree of confidence.

This work has multiple ramifications for our immunological understanding and future clinical applications. Firstly, our data challenge the dogma that CMV infection has solely deleterious effects on immunity. We find that CMV polarisation of T cell immunity towards a more primed and effector footing is associated with improved immune surveillance and tumour rejection. Our data also show that CMV buffers the effect of ageing in MM, with increasing age only associated with an increased hazard for death once CMV serostatus has been controlled for.

Secondly, this study has important clinical implications as it suggests that patients seronegative for CMV are at risk of poorer outcomes if not treated with cICB, yet are also at increased risk of high grade irAEs. CMV seronegative patients may therefore particularly benefit from irAE mitigation strategies. Whilst CMV may similarly interact with the response of other tumour types to ICB, the previous largest reported study in lung cancer did not find a direct association of seropositivity with OS however [50] and further studies of the effect of CMV and other CVIs on the responses of other immunosensitive cancers to ICB are vital. Finally, as immunotherapies become increasingly complex and costly, we show that non-specific immune stimulation from CVI associates with substantial epidemiological and clinical impact in melanoma. The identification of *TBX21*, encoding T-bet, as a master transcription factor determining the benefit associated with CMV seropositivity delineates a molecular pathway that may be amenable to intervention to enhance ICB efficacy. Our findings also have implications for trial design of cancer vaccines where it will be important to dissect benefits brought by targeted agents from those conferred by generic adjuvants. As such, this work reinforces the central importance of factors that influence systemic immunity in the development of metastatic cancer and response to immunotherapy.

## Methods

### Patients

Recruitment occurred between 23 November 2015 and 8 July 2024. Patients eligible for ICB in the Oxford University Hospitals NHS Foundation Trust were recruited prospectively. These included patients with advanced or metastatic melanoma (MM), renal cell carcinoma (RCC), mismatch repair deficient colorectal cancer (CRC), mesothelioma (meso), and cutaneous squamous cell carcinoma (cSCC). Patients due to receive adjuvant ICB for MM and RCC were also eligible. Informed consent was given by all patients to donate samples to the Oxford Radcliffe Biobank (Oxford Centre for Histopathology Research ethical approval reference 19/SC/0173, project nos. 16/A019, 18/A064 and 19/A114) and grant access to their routine clinical data. There was no compensation for this consent. Immunological and clinical outcome analyses were confined to MM patients who were treated with either cICB (ipilimumab 3mg kg^−1^ q3w plus nivolumab 1mg kg^−1^ q3w for 4 cycles followed by maintenance nivolumab) or sICB (pembrolizumab 200mg q3w or 400mg q6w, or nivolumab 240mg q2w or 480mg q4w).

(OxCITE cohort, n= 396, median follow up 40.1 months) (Metastatic melanoma cohort, n = 304, median follow up 55.8 months) (CMV seropositive MM cohort, n = 126, median follow up 46.9 months) (CMV seronegative MM cohort, n = 147, median follow up 53.4 months)

### Clinical outcomes

Patient demographic, clinical, haematological, biochemical and radiological data were extracted from electronic patient records. All patients who received at least 1 cycle of ICB were included in the analysis. irAEs were assessed according to the National Cancer Institute’s Common Terminology Criteria for Adverse Events, version 5.0. PFS was determined as time from treatment initiation to clinical or radiological evidence of progression or death. OS was defined as time from treatment initiation to death from any cause.

### Sample collection

Up to 50ml of whole blood was collected from each patient, in EDTA vacutainer tubes, immediately prior to ICB administration. Plasma and PBMCs were separated immediately by density centrifugation using Ficoll-Paque. Cell-subset sorting for bulk RNA-seq was performed using Miltenyi Biotec magnetic separation, with positive selection for CD8^+^ T-cells performed per manufacturer instructions at 4°C. Baseline full blood count data were obtained from electronic patient records, generated via Sysmex XN series analyzers (Sysmex corporation). The blood sample taken closest to initiation of ICB, between day –30 and day 0, was used for analysis. CMV IgG antibody titres were measured using patient plasma diluted 1 in 2 from day 0 samples on an Abbott architect i2000 following GLP within the John Radcliffe hospital, Oxford. Median value for CMV negative patients =0.2 AU/ml (mean 0.3 AU/ml, IQR: 0.1-0.3 AU/ml). For CMV positive patients saturating values (where AU*>*250 AU/ml) were read as 250 AU/ml, median value = 87.5 AU/ml (mean 105.9 AU/ml, IQR: 53-156.8 AU/ml).

### Flow cytometry

Cryopreserved patient PBMC samples were thawed at 37°C and washed with HBSS. 1×10^6^ PBMCs were plated, and viability staining was performed with Near IR Fixable Viability Stain (ThermoFisher, L34975) or Fixability Viability Stain 440UV (BD Biosciences, for 30min at 4°C. PBMCs were washed with HBSS supplemented with 5% FCS and BD Horizon Brilliant Stain Buffer (BD Biosciences, 563794). Staining for T cell surface markers was performed for 30min at 4°C using antibodies directed against surface markers listed in Supplementary Table 5. PBMCs were washed once in cell staining buffer (BioLegend, 420201) and incubated with Streptavidin (BV785, BioLegend 405249) for 30min at 4°C for IgG4 staining. Following surface staining, PBMCs were permeabilized with Foxp3/Transcription Factor Staining Buffer Set (Invitrogen, 00-5523-00) for intra-nuclear staining or Transcription Factor Buffer Set (BD Biosciences, 562574) for cytoplasmic staining, for 30min and incubated with intracellular staining antibodies listed in Supplementary Table 5. PBMCs were resuspended in 2% paraformaldehyde and acquisition was performed on a FACSSymphony A5 Cell Analyser (BD Bioscience) or a Fortessa X-20. Data was analysed using FlowJo v10.7.1 (BD Biosciences).

### Nucleic acid extraction

Post-selection cells were spun down at 4°C and resuspended in 350µl of RLTplusbuffer with 1% DTT and transferred to 1.5ml Eppendorf tubes. Samples were stored at –80°C for batched RNA extraction. Homogenisation of the sample was carried out using the QIAshredder kit (Qiagen). The AllPrep DNA/RNA/miRNA kit (Qiagen) was used for RNA extraction. DNase I was used during the extraction protocol to minimise DNA contamination. RNA was eluted into 35µl of RNase-free water. The amount of RNA present was quantified by Qubit analysis, and RNA samples were stored at –80°C until sequencing.

### Bulk RNA-seq preparation and analysis

RNA was thawed on ice prior to mRNA isolation using the the NEBNext Poly (A) mRNA Magnetic Isolation Module Kits. Poly(A) RNA was either sequenced using 150-base pair-end sequencing on an Illumina NovaSeq or using 75-base pair-end sequencing on an Illumina HiSeq-4000, both at the Oxford Genome Centre. Reads were aligned to CRGh38/hg38 using HISAT2 and read count data was generated using HTSeq. High-mapping quality reads were selected based on MAPQ score using bamtools. Marking and removal of duplicate reads were performed using picard (v.1.105) and samtools was used to pass through the mapped reads and calculate statistics. A total of 217 (186 with CMV serology, 82 CMV+ and 104 CMV-) high quality baseline CD8^+^ T cell transcriptomes were included in the analysis along with 164 (145 with CMV serology, 68 CMV+ and 77 CMV-) day 21 transcriptomes, with 156 (137 with CMV serology, 64 CMV+ and 73 CMV-) paired samples. Normalised counts were produced using DESeq2 (v1.40.1).

DESeq2 was used to perform differential expression analysis at baseline to identify genes modulated by CMV. Age, sex and sequencing batch were corrected for in the design formula and only genes with a mean count *>*10 were included in the analysis. Induced and downregulated genes were treated separately for pathway analysis using the R package XGR (v1.1.9). The GOBP database was used with all annotated genes (n= 14665) serving as a background. The ontology algorithm was specified as “elim” along with an elimination P value of 0.01, and a hypergeometric test was employed.

CMV score was determined using the normalised counts of differentially expressed genes at baseline between CMV seropositive and seronegative patients. Gene counts were isolated for baseline and day 21 samples and were incorporated in a principal component analysis (PCA) using the R package stats. The first principal component was extracted and was used in a ROC analysis (using the R package pROC v1.18.2) to predict CMV serostatus. The number of genes to be included in the CMV score was determined through iterative PCA and ROC analyses between an adjusted p-value threshold, obtained from the DESeq2 results, of 0.05-10^−15^ (encompassing 7175 to 44 genes). The threshold yielding the maximum area under the curve (0.90) was selected, including 103 genes. Cytotoxicity score was determined using a set of 50 genes that had been previously found to correlate with *IFNG* expression (Supplementary Table 3). The normalised counts of this gene set were extracted from baseline and day 21 samples and the score was calculated using a geometric mean.

Transcription factor (TF) analysis was performed in R using decoupleR (v2.6.0). The network from OmniPath, obtained for “human” and without splitting complexes, was filtered to contain TFs that were detected in the CD8+ T cell count matrix, yielding 792 testable transcription factors. Counts data was normalised using variance stabilising transform in DESeq2. Normalised counts data along with differentially expressed genes (FDR *<*0.05) according to CMV serostatus at baseline were used in a univariate linear model for TF activity inference. TFs with induced acitivity in CMV seropositivity with nominal (P*<*0.05) significance were included in downstream analysis.

### Adaptive receptor analysis

MiXCR was used to map bulk CD8^+^ reads on reference sequences of V, D and J genes, and to quantitate TCR clonotypes from mapped reads using complementarity-determining region 3 (CDR3) gene regions. The non-default partial alignments option (OallowPartialAlignments = true) was applied to preserve partial alignments for the assembly step. Three iterations of read assembly were performed to increase the number of assembled reads containing the CDR3 region using assemblePartial action. Position quality scores were used to switch on the frequency-based correction of clonotype assembling and clustering (ObadQualityThreshold = 15). Clones were called according to the nucleotide sequence and a total of 591781 *TRA* chains (median per sample = 1191 chains, range = 152-4837 chains) and 1008395 *TRB* chains (median per sample = 2068 chains, range = 260-9430 chains) were identified. Clone size was determined for *TRB* chains, with large clones being defined as those with a peripheral repertoire occupancy (determined by cloneCount/ Total *TRB* count) *>*0.5% while intermediate and small clones were those found to occupy *>*0.05% and *<*0.05% of the repertoire, respectively.

### scRNA-seq preparation and analysis

A combination of fresh (8 samples) and thawed (28 samples) positively selected CD8^+^ T cells were counted and oil-partitioned into single cell droplets, followed by cell lysis and reverse transcription on a 10X Genomics Chromium System. A total of 6000 cells were loaded onto each partitioning reaction. 5’ GEX and V(D)J libraries were constructed from cDNA using the Chromium Next GEM Single Cell 5’ Reagent Kit v2 (Dual Index). Droplet generation and reverse transcription were performed on individual samples, but sample generation and sequencing were performed in batches. Sequencing was performed on an Illumina HiSeq4000: 75 bp paired-end (PE) reads for 5’ RNA libraries, 150 bp PE reads for the V(D)J libraries to a depth of about 20,000-50,000 reads per cell.

FASTQ files were generated from raw Illumina BCL outputs using Cellranger (v6.0.1) for GEX libraries and Cellranger VDJ for V(D)J libraries. For QC, the R package scater was used to identify single-cell outliers and the R package scran was used to identify doublets. Post-initial pre-processing, CD8+ T cells were filtered for expression of *CD3D* and either *CD8A* or *CD8B* along with absence of *CD14* expression. Seurat was used for further QC, read normalization and identification of variable features. Cells used in analysis were those with *>*500 total UMIs, *>*300 features, and mitochondrial gene percentage *<*20%. Genes expressed in *<*5 cells were also excluded. Principal component analysis (PCA) was performed on scaled data, with the top 16 principal components being selected for dimensionality reduction and uniform manifold approximation and projection (UMAP) clustering. Cell clusters were generated between resolutions of 0.1 and 2.0 using the R package clustree, with values of 1.5-2.0 demonstrating high cluster stability. Cells were assigned to subsets based on curated transcripts per million normalized counts for CD8^+^ T cells from previous public T cell cancer scRNAseq datasets using SingleR [4, 51–54].

*TBX21* expression was imputed using the MAGIC package (v2.0.3.999) [55]. Subset expression of *TBX21* was plotted using the ggridges (v0.5.4) package, and the effector subset was found to have the highest median imputed expression (0.44 units), and they were therefore selected for downstream analysis. Clones were defined as those with a detected CDR3 *TRA* and/or *TRB*. Median imputed TBX21 was determined per effector clone for 18 anti-PD-1 treated patients at baseline and day 21. 1461 (median per sample = 78 clones) clones from baseline samples and 1581 clones from day 21 samples (median per sample = 66 clones). Clone size was determined using the same criteria as for bulk RNA-seq.

### Statistical analysis

Statistical analyses were performed using R version 4.4.0. Survival analyses were performed using the survival (v3.7-0) and survminer (v0.4.9) packages, while cumulative incidence and competing risks regression using tidycmprsk (v1.0.0). Plots were generated using ggplot2 (v3.5.1). Unless stated otherwise, all Cox proportional hazards, cumulative risk, and logistic regression analyses were performed adjusting for age, sex, *BRAF* status, previous *BRAF* inhibitor therapy, and single versus combination ICB. Median follow-up time was calculated using the reverse Kaplan-Meier method described by Schemper and Smith[56]. For survival analysis, patients with a minimum of 6 months follow-up were included. Survival analysis was censored at 90 months post-initiation of systemic treatment. CMV effect on PFS and OS was assessed separately for patients with metastatic melanoma receiving sICB or cICB, initially via Kaplan-Meier curves with univariate log-rank tests then multivariable Cox proportional hazards models. Univariate cumulative incidence estimates were calculated for time to irAEs using a semi-competing risks model, with death as the competing risk. Time to first irAE of Grade 1-2 irAE and first Grade 3+ irAE were assessed separately for their association with CMV status. Significance testing was performed using Gray’s test[57]. Competing risk regression was then used to calculate the Fine-Gray subdistribution hazard for association of CMV status and each irAE measure[58]. Adjusted odds ratios for irAEs were calculated using binomial logistic regression correcting for ICB type, age, sex, *BRAF* mutation status and prior BRAF inhibition. The impact of irAEs on PFS and OS was assessed using a time-dependent Cox-regression model to account for guarantee-time bias[59]. First, Grade 1-2 and Grade 3+ irAEs were considered as time-dependent covariates. Second, organ-specific irAEs were assessed, again as time-dependent covariates, adjusting for each different irAE and the same covariates used in the binary logistic regression analysis.

Correlation analysis was performed using a Spearman’s Rank Correlation test and distributions were compared using either a Wilcoxon signed rank test, for paired data, or a Wilcoxon rank sum test, for unpaired data. Odds ratios (ORs) were generated using a Fisher’s exact test, while adjusted ORs were generated using a generalised linear model (family = “binomial”). Age and gender matching between the OxCITE cohort and the UKB was performed using the R package MatchIt (v4.5.5). Permutation testing was performed using a two-tailed alternative hypothesis and 10,000 Monte-Carlo replicates, using the R package coin (v1.4-2). The matching process was bootstrapped 1000 times and the median P value was taken.

## Data Availability

All data produced in the present study are available upon reasonable request to the authors – they will be made available with scripts via codecapsule

## Acknowledgements

We are grateful to all patients who contributed samples and participated in the study without compensation. We thank all the staff of the Day Treatment Unit, Oxford Cancer Centre and the Brodey Centre at the Horton General Hospital. We are grateful to all the staff of the Oxford University Hospitals NHS Foundation Trust haematology, microbiology and biochemistry laboratories and thank T. James for his facilitation, as well as the staff of the Oxford Radcliffe Biobank and Churchill Hospital Sample Handling Lab.

## Declarations

- Funding B.P.F. is funded by a Wellcome Career Development Award (226535/Z/22/Z) and the study was previously funded by a Wellcome Intermediate Clinical Fellowship to B.P.F. (201488/Z/16/Z). P.K. is funded by a Wellcome Discovery Award 222426/Z/21/Z and P.K. and B.P.F. are recipients of a CRUK programme grant DRCNPG-Nov22/100005 ‘Defining the role of MAIT cells in Cancer Immunotherapy’. G.M. is a DPhil student whose fees have been covered by Etcembly. J.J.G. is supported by an NIHR Clinical Lectureship. R.A.W. is funded by the CRIS cancer foundation postdoctoral fellowship (BBD00040), previously by a Wellcome Trust Doctoral Training Fellowship (no. BST00070). The Oxford Radcliffe Biobank and Oxford Centre for Histopathology Research are supported by the University of Oxford, the Oxford CRUK Cancer Centre and the NIHR Oxford Biomedical Research Centre (Molecular Diagnostics Theme/Multimodal Pathology Subtheme), and the NIHR Cancer Research Network (CRN) Thames Valley network. M.R.M. and B.P.F. are supported by the NIHR Oxford Biomedical Research Centre. The views expressed are those of the authors and not necessarily those of the NHS, the NIHR or the Department of Health.
- Conflict of interest in the last three years BPF has performed consultancy for NICE Consultancy, Roche, Pathios, UCB, TCypher and been given speaker fees by GSK, UCB, BMS, all of which is outside the submitted work. MRM reports grants from Roche, grants from Astrazeneca, grants from GSK, other from Novartis, grants and other from Immunocore, other from BMS, other from Pfizer, other from Merck/MSD, other from Regeneron, other from BiolineRx, other from Replimune, grants from GRAIL; all outside the submitted work.
- Ethics approval and consent to participate Informed consent was given by all patients to donate samples to the Oxford Radcliffe Biobank (Oxford Centre for Histopathology Research ethical approval reference 19/SC/0173, project nos. 16/A019, 18/A064 and 19/A114) and grant access to their routine clinical data. There was no compensation for this consent.
- Materials availability Not applicable.
  - Data availability Raw sequencing data will be made available via a Data Access Arrangement between applicants and the University of Oxford. Normalised gene expression matrices with associated minimal anonymised patient information data (covariates required to perform analyses in paper) will be available upon publication via Oxford Research Archive (https://ora.ox.ac.uk).
  - Code availability Code to perform all analyses and make figures is provided on Fairfax lab Git Hub
  - Author contribution B.P.F. conceptualised and oversaw the study. G.M. and M.L. performed primary data analyses, with assistance from R.A.W, B.S., J.J.G, I.N. G.M. constructed figures. R.A.W. performed scRNA-seq experiments and analyses. N.C., M.J.P., M.R.M., B.P.F., V.W., M.L. and R.A.W. recruited patient participants. C.A.T., D.M., O.T., R.C., O.A., J.B., P.K.S., S.M., S.D., F.M.S, A.V.A. collected and processed blood samples. S.M., D.M., F.M.S, A.V.A. generated RNA-seq libraries. R.A.W., M.L. and B.P.F. collected clinical follow-up data. C.A.T., O.T., D.M., S.M., performed flow cytometry experiments. R.A.W., O.T., W.Y., J.J.G. and B.P.F. performed data analysis. M.R.M., M.Y., S.D. provided infrastructure support. A.M., A.Y.C and E.N., provided support with analyses of UKB CMV data. P.K. provided immunological support and oversight re. CMV. B.P.F. wrote the manuscript with input from M.L., G.M., R.A.W., P.K.. All authors read, edited and approved the final manuscript.

## Supplementary Tables

- Supplementary Table 1 CD8^+^ T cell CMV differentially expressed genes
- Supplementary Table 2 GOBP pathway analysis of CD8^+^ T cell CMV differentially expressed genes
- Supplementary Table 3 Genes incorporated into Cytotoxicity score
- Supplementary Table 4 CMV associated transcription factors – output from decoupleR
- Supplementary Table 5 Antibodies utilised in flow cytometry

## Supplementary Data

- Supplementary Figure 1 Flow gating strategy

## Extended Figures

**Fig. S1.**
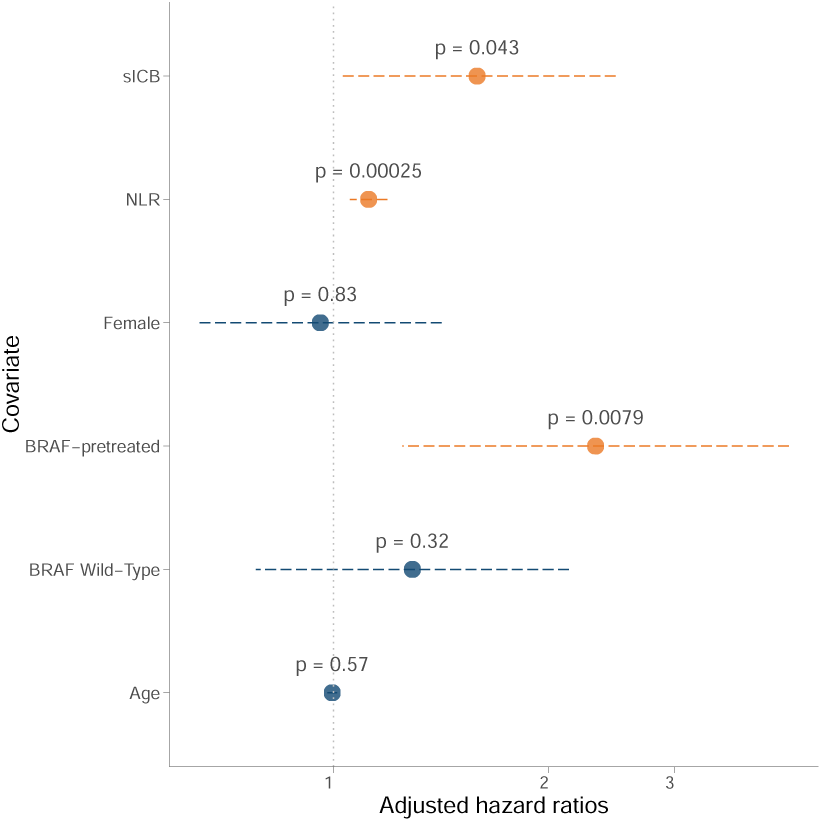
Cox proportional hazards model of covariates including neutrophil:lymphocyte ratio and occurrence of death in MM patients

**Fig. S2.**
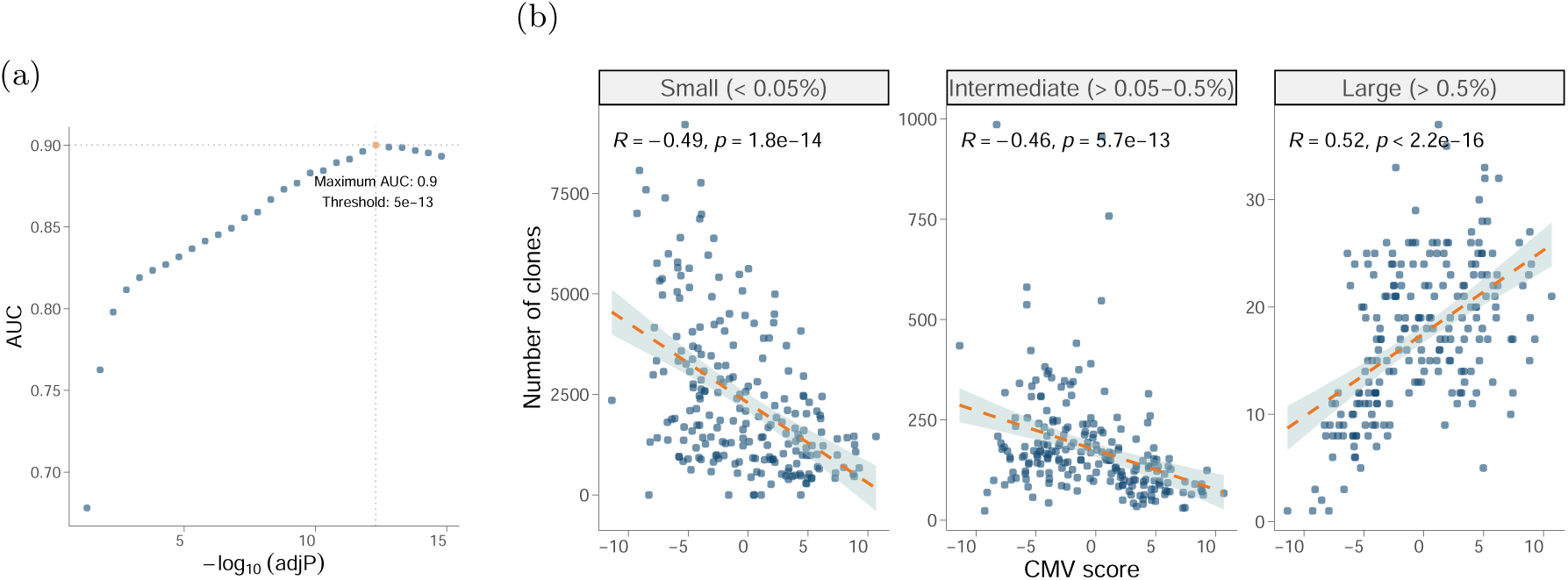
(a) ROC analysis of CMV seropositivity prediction using the first principal component of DEGs between baseline CD8^+^ samples based on CMV status. Area under the curve (y axis) is plotted against the –log10(adjusted P value) threshold (x axis) used to determine genes to be incorporated in the score with the optimal threshold labelled (b) Correlation of CMV score with number of CD8^+^ clones stratified by repertoire occupancy in baseline samples (Spearman’s Rank Correlation test)

**Fig. S3.**
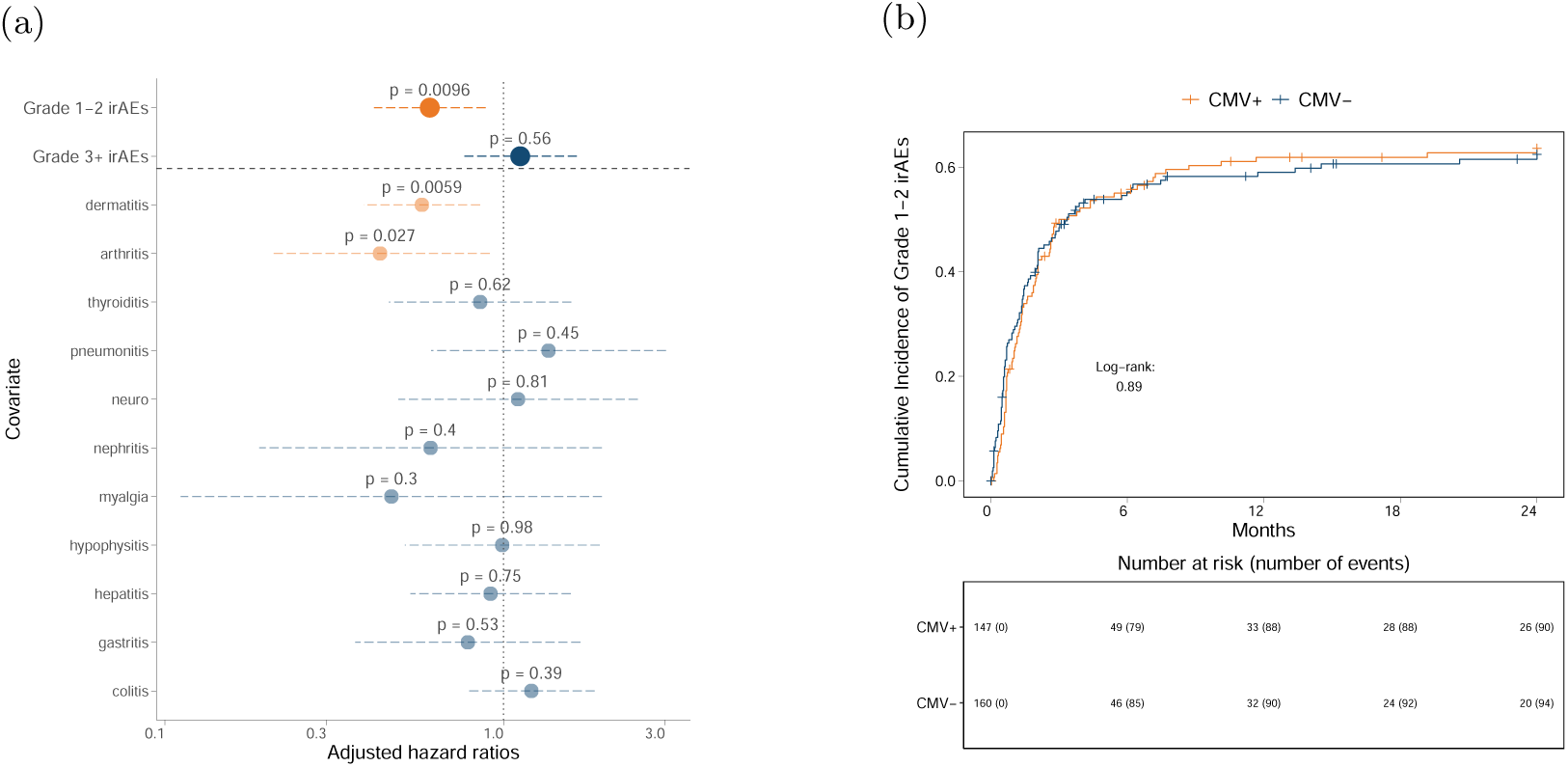
(a) Cox proportional hazards analysis of irAEs by grade and by organ specificity demonstrates grade 1-2 irAEs and specifically dermatitis and arthritis bestow reduced risk of death (b) Cumulative incidence of grade 1-2 irAEs according to CMV status

**Fig. S4.**
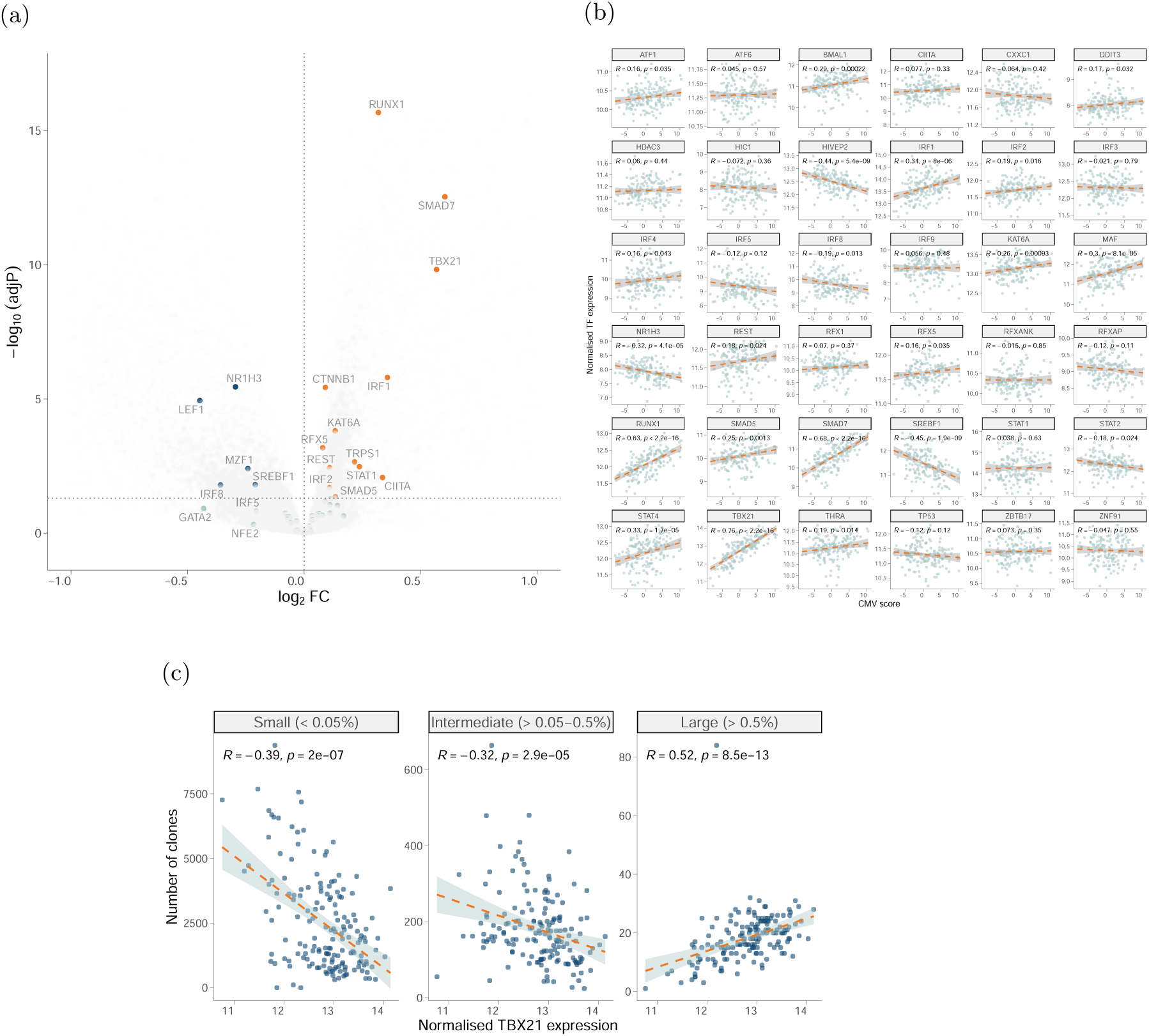
(a) Differential expression of transcription factors with altered activity in CMV seropositivity in baseline CD8^+^ samples (b) Correlation of transcription factors with induced activity in CMV seropositivity with CMV score (Spearman’s Rank Correlation test) (c) Baseline CD8^+^ *TBX21* correlation with CMV score (Spearman’s Rank Correlation test)

**Fig. S5.**
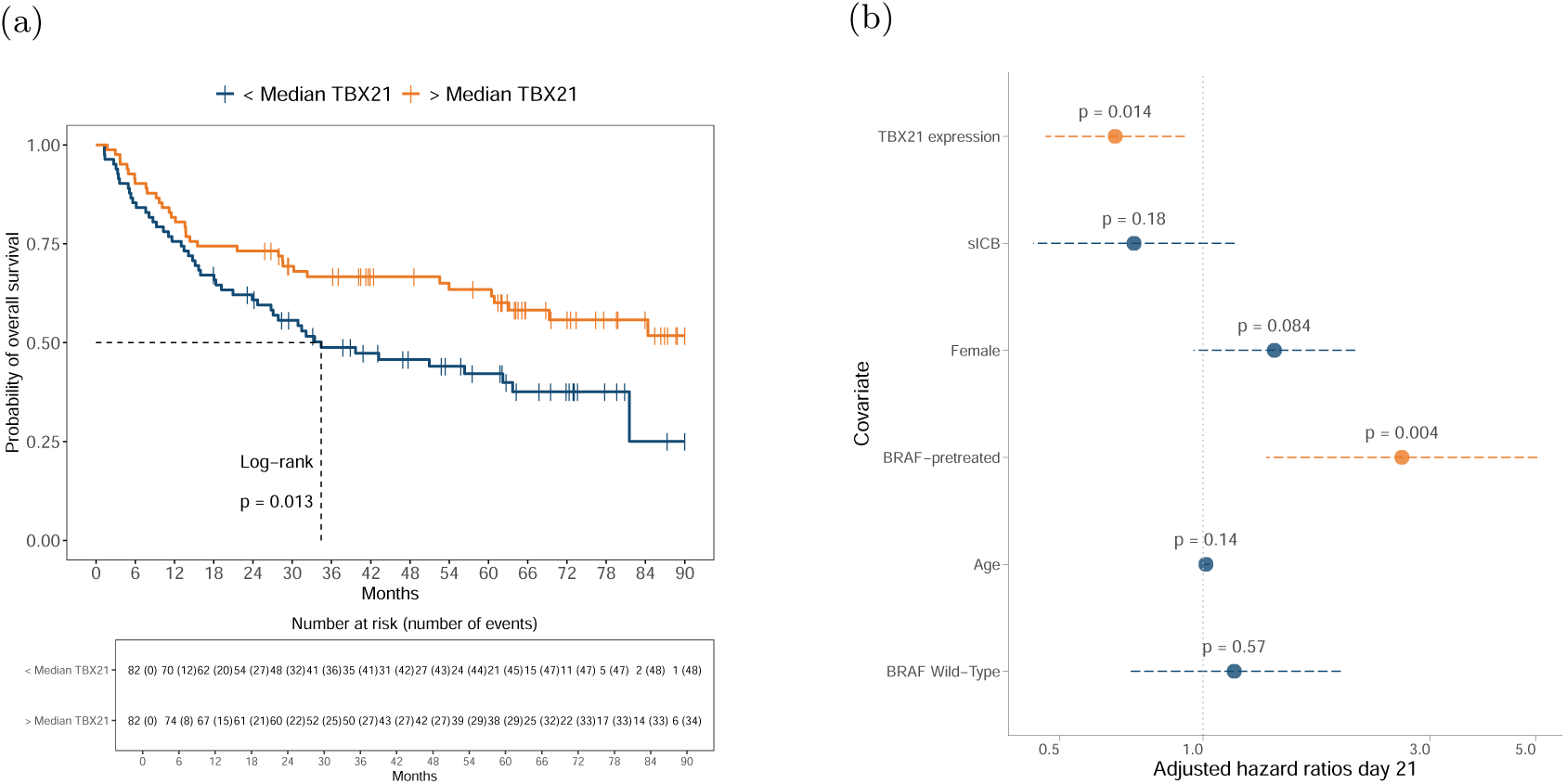
(a) Kaplan Meier analysis demonstrates significantly improved in OS in patients with above median *TBX21* post one cycle of immunotherapy (P=0.013, log-rank test) (b) Cox proportional hazards model indicates that increased post-treatment *TBX21* is associated with improved PFS when adjusting for ICB type, age, sex, *BRAF* mutational status and prior *BRAF* inhibition

**Fig. S6.**
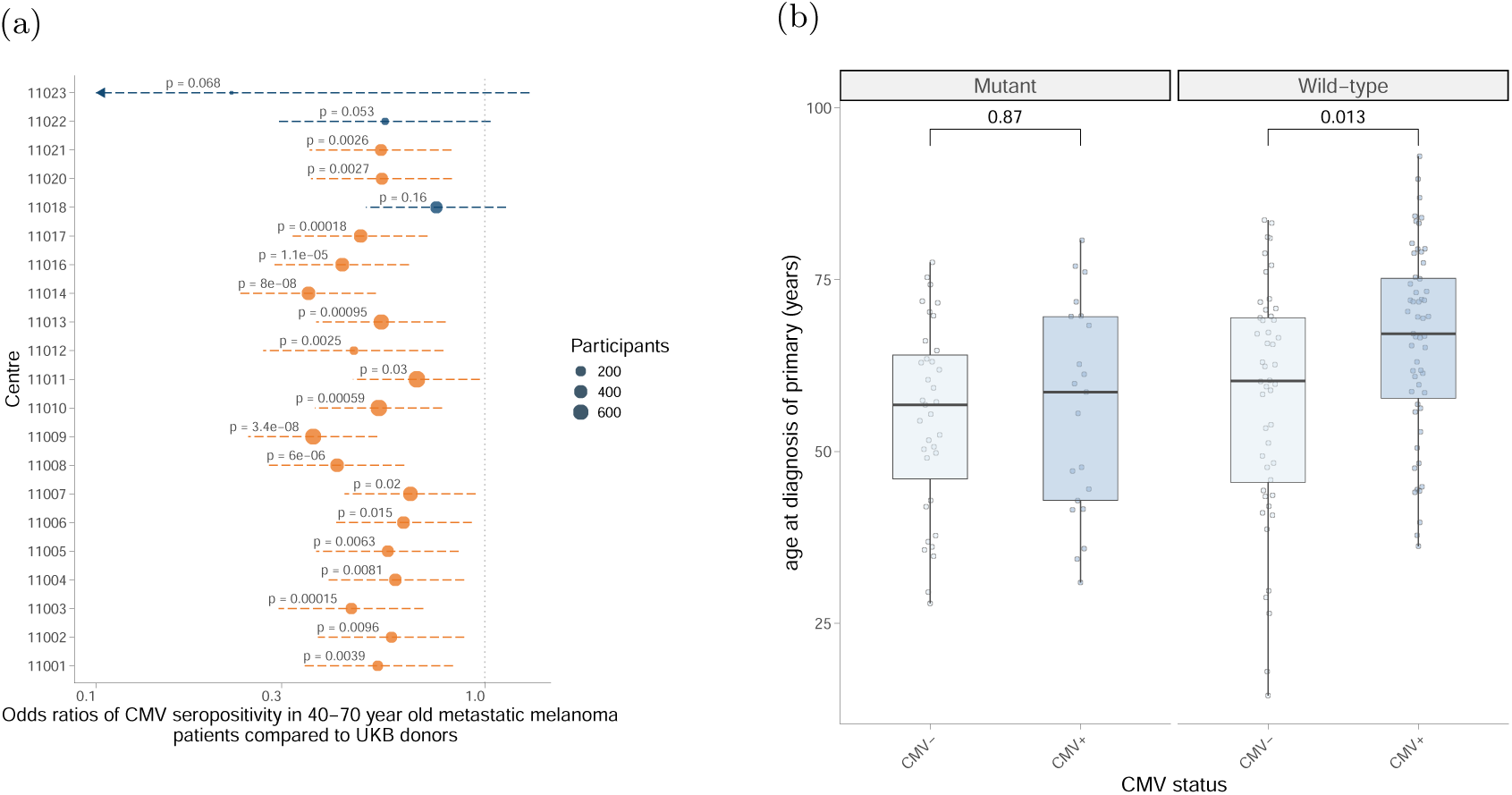
(a) Relative depletion of CMV seropositive patients in MM cohort versus UKBB data according to recruitment centre (b) Age of diagnosis of primary melanoma by CMV serotype (CMV serology determined at diagnosis of metastatic disease)

